# Multiomic Mendelian randomization-based insights into the role of neutrophil extracellular trap-related genes in sepsis

**DOI:** 10.1101/2024.12.24.24319599

**Authors:** Yanbo Liu, Yuhui Li, Jinmin Chen, Yongxia Cai, Lukai Lv

## Abstract

Sepsis is a life-threatening organ dysfunction caused by a dysregulated host response to infection. Neutrophil extracellular traps (NETs) have been implicated in the pathogenesis of sepsis, yet the precise role of NET-related genes (NRGs) remains unclear. This study employed a multiomic Mendelian randomization (MR) approach, leveraging genetic variants as instrumental variables to investigate the relationships between NRGs and sepsis risk. We systematically identified 69 NRGs based on literature and database reviews. Utilizing the IEU OpenGWAS Project database, we extracted genetic data for sepsis cases and controls. Expression quantitative trait loci, methylation quantitative trait loci (mQTLs), and protein quantitative trait loci (pQTLs) associated with NRGs were obtained from the eQTLGen Consortium, mQTL meta-analysis, and deCODE Genetics datasets, respectively. We employed the inverse variance-weighted method, supplemented by MR-Egger regression, weighted median, and Bayesian colocalization analysis, and identified four genes (*CXCR1*, *CXCR2*, *ENTPD4,* and *MAPK3*) significantly associated with sepsis risk. Three CpG sites associated with these genes were identified through mQTL-based MR analysis. Additionally, ten proteins showed significant associations with sepsis risk in pQTL-based MR analysis. Summary-data-based MR and colocalization analyses confirmed the causal relationship between *CXCR2* and sepsis, which remained unaffected by pleiotropy. The DNA methylation level at cg06547715, located in the *CXCR2* enhancer region, was inversely correlated with *CXCR2* expression and sepsis risk. These findings suggest that NRGs, particularly *CXCR2*, play a crucial role in sepsis susceptibility and that the DNA methylation status of *CXCR2* may modulate gene expression, influencing sepsis risk. This study provides novel insights into the molecular epidemiology of sepsis and highlights the potential of NRGs as therapeutic targets. Targeting *CXCR2* and its regulatory mechanisms may offer a new avenue for sepsis management. These findings contribute to the theoretical understanding of sepsis pathogenesis and pave the way for future research into precision medicine for sepsis.

## Introduction

Sepsis, an abnormal response to infection, is a leading cause of death among critically ill patients worldwide[1, 2]. Marked by systemic inflammation, it can quickly lead to organ failure and death in the absence of prompt treatment[3, 4]. The pathophysiology of sepsis is complex, involving an intricate interplay among the host immune system, coagulation cascade, and endothelium[5, 6]. Despite advances in critical care and antibiotic therapy, sepsis management continues to pose a significant challenge, highlighting the need for attaining a deeper understanding of the mechanisms underlying this condition and for the development of novel therapeutic strategies[7–9].

Recent studies have highlighted the role of neutrophil extracellular traps (NETs) in the pathogenesis of sepsis, particularly in the context of COVID-19 infections, wherein NETs have been implicated in the hyperinflammatory response and microthrombi formation[10, 11]. Neutrophils, the most abundant leukocytes in the human body, play a pivotal role in the innate immune response against infections[12, 13]. Among their various defense mechanisms, the formation of NETs has emerged as a critical process in pathogen clearance[14]. NETs are web-like structures composed of DNA, histones, and antimicrobial proteins, which are released by neutrophils through a process termed NETosis[15, 16]. While NETs are effective in trapping and neutralizing a broad spectrum of microbes, excessive NET formation has been implicated in the pathogenesis of various inflammatory and thrombotic conditions, including sepsis[17, 18]. The recognition of NETs in the context of sepsis has led to a paradigm shift in understanding the role of neutrophils in disease pathogenesis. NETs have been shown to contribute to the formation of microthrombi, exacerbating tissue injury and organ dysfunction during sepsis[19, 20]. Furthermore, NETs can serve as a source of proinflammatory mediators, potentially driving the systemic inflammation observed in sepsis[21, 22].

The involvement of NETs in sepsis is increasingly being recognized, but the mechanisms governing their formation and their impact on disease severity remain poorly understood[23]. Given the potential impact of NETs on sepsis outcomes, elucidating the genetic and molecular factors that regulate NETosis is an urgent need. Identifying key regulators of NET formation may reveal novel therapeutic targets for mitigating the detrimental effects of excessive NET production in sepsis. In this study, we employed two-sample Mendelian randomization (MR), a powerful epidemiological tool that leverages genetic variants as instrumental variables (IVs)[24]. Through the analysis of large-scale genome-wide association studies (GWASs) and quantitative trait loci (QTL), we investigated the causal relationship between NETosis-related genes (NRGs) and sepsis risk.

## Materials and methods

### Data acquisition

#### Identification of NRGs

We systematically identified and curated a comprehensive list of 69 NRGs based on a thorough review of the existing literature and databases[14, 25], focusing on the role of NETs in immunity and various diseases. The selection criteria for these genes included their involvement in the formation, regulation, and biological impact of NETs, encompassing ligands and receptors that stimulate NETs, downstream signaling pathways, and molecules known to be components of the NET framework (**S1 Table**).

#### Data sources of methylation, expression, and protein QTLs

We accessed publicly available genomic data relevant to sepsis using the “IEU OpenGWAS Project” database. This database, known for its extensive collection of GWAS data, provided a robust platform for our investigation. Specifically, we extracted data from the hospital episode statistics, which included two sepsis datasets—a primary dataset (ieu-b-4980) consisting of 11,643 sepsis cases and 474,841 control participants, and a validation dataset (ieu-b-5088) comprising 11,568 sepsis cases and 451,301 control subjects (**S2 Table**). Both datasets included individuals of European ancestry, ensuring genetic homogeneity was maintained for the MR analyses. To screen for expression quantitative trait locus (eQTL) IVs of the NRGs, genetic variants robustly associated with gene expression within a 1000 kb range on either side of the coding sequence (cis) were extracted from the eQTL summary statistics obtained from the eQTLGen Consortium (https://www.eqtlgen.org/cis-eqtls.html). The eQTLGen Consortium includes information on 10,317 single nucleotide polymorphisms (SNPs) associated with traits from 31,684 individuals. Using the methylation quantitative trait locus (mQTL) data from a meta-analysis of summary statistics from two cohorts (n = 1980) available at (https://yanglab.westlake.edu.cn/data/SMR/LBC_BSGS_meta_lite.tar.gz), cis-mQTL IVs robustly associated with NRG methylation were extracted. From the deCODE Genetics dataset of 4907 proteins (https://www.decode.com/summarydata/), cis-protein quantitative trait locus (pQTL) IVs associated with the expression of proteins related to NRGs were selected; all SNPs included in the initial analysis met the criterion of *P* < 5 × 10^−8^.

### MR analysis

#### Assumptions and approach

We used two-sample MR to examine the causal relationships between the QTLs of 69 NRGs and sepsis development. The two-sample MR technique utilizes genetic variants as IVs to estimate the causal effects of modifiable risk factors by leveraging the random assortment of alleles at conception. The application of MR in this study was based on three key assumptions—(1) IVs are strongly associated with the exposure of interest; (2) IVs are independent of confounders that could influence the outcome; (3) IVs affect the outcome solely through exposure. Deviations from these assumptions, particularly through pleiotropy, where IVs affect the outcome through multiple biological pathways, can bias MR estimates.

#### IV selection

The selection of IVs for each NRG was based on established criteria. eQTLs, mQTLs, and pQTLs associated with NRGs were considered as potential IVs. The criteria for IV selection included the following: (1) a genome-wide significant association between the SNP and the gene (*P* < 5 × 10^−8^); (2) minimal linkage disequilibrium among SNPs (r^2^ < 0.1) with a threshold distance of 10,000 kb to ensure independence; (3) exclusion of SNPs with palindromic sequences to harmonize exposure and outcome datasets, followed by MR-PRESSO testing to exclude outlier SNPs; (4) an F-statistic ≥ 10 [F-statistic = (beta/se)^2^][26] to ensure sufficient statistical power for the IVs in the analysis.

#### Two-sample MR analysis and assessment of heterogeneity and pleiotropy

The primary two-sample MR analysis was conducted using the inverse variance-weighted (IVW) method. Before IVW analysis, the datasets were assessed for heterogeneity using Cochran’s Q test with a significance threshold of *P* < 0.05. A random-effects IVW model was used in the presence of significant heterogeneity, while a fixed-effects IVW model was applied when no heterogeneity was detected. Complementary MR methods were employed to strengthen the findings, including MR-Egger regression, weighted median, weighted model, and simple model. The *P-*values were adjusted to control the false discovery rate (FDR) at α = 0.05 using the Benjamini-Hochberg method. The estimated causal effects were quantified using the odds ratio (OR) and the 95% confidence interval (CI). The MR-Egger regression intercept test was used to assess potential pleiotropy, with a non-significant *P* > 0.05 indicating the absence of horizontal pleiotropy. Cochran’s Q statistic from both the MR-Egger and IVW methods was used to detect heterogeneity among various causal effects, with a Cochran’s Q test *P*<0.05 indicating heterogeneity[26, 27]. We also employed the Steiger filtering method to verify the directional associations among eQTLs, mQTLs, and sepsis.

### Summary-data-based MR (SMR)

#### SMR analysis and heterogeneity in the dependent instrument (HEIDI) testing

SMR was employed to ascertain the causal inferences of NRG methylation, expression, and protein abundance with the risk of sepsis, based on the top associated cis-QTLs. SMR can reach a much higher statistical power than conventional MR analysis when exposure and outcome are available from two independent samples with large sample sizes. The top associated cis-QTLs were selected by considering a window centered around the corresponding gene (±1000 kb) and passing a *P*-value threshold of 5.0 × 10^−8^. The SNPs with allele frequency differences larger than the specified threshold (set as 0.2 in the present study) between any pairwise datasets, including the linkage disequilibrium reference sample, the QTL summary data, and the outcome summary data, were excluded. The HEIDI test was applied to distinguish pleiotropy from linkage, where P-HEIDI values <0.01 were considered to be likely due to pleiotropy and thus discarded from the analysis. SMR and HEIDI tests were implemented using the SMR software (version 1.3.1).

#### Colocalization analysis

To confirm the results, we used the coloc R package (version 5.2.3, available at https://chr1swallace.github.io/coloc/)[28] for Bayesian collocation analysis to estimate the posterior probabilities of shared variants. The collocation analysis relies on four hypotheses—(1) Phenotype 1 (GWAS) and Phenotype 2 (e.g., eQTL) show no significant SNP associations in a specific genomic region (H0); (2) Phenotype 1 (GWAS) or Phenotype 2 (e.g., eQTL) shows a significant association with SNPs in a specific genomic region (H1/H2); (3) Phenotype 1 (GWAS) and Phenotype 2 (e.g., eQTL) are significantly associated with SNPs in a specific genomic region, but driven by different causal variants (H3); (4) Phenotype 1 (GWAS) and Phenotype 2 (e.g., eQTL) are significantly associated with SNPs in a specific genomic region and driven by the same causal variant (H4). We extracted all SNPs within 100 kb upstream and downstream of the lead SNP in the GWAS for collocation analysis and calculated the posterior probability H4 (PPH4). In academic contexts, a PPH4 value exceeding 0.75 is conventionally employed as the threshold to indicate evidence of collocation between GWAS and QTL associations.

### Ethical Considerations

The data sources utilized in this study, including the IEU OpenGWAS Project, eQTLgen, the mQTL summary data of McRae et al., and the deCODE Genetics dataset, are all publicly accessible and do not contain any personal identifiers or sensitive personal information that would require consent.The study followed the Declaration of Helsinki principles and human subject research guidelines.

## Results

### Analytical process overview

In this study, two-sample MR analysis of IVs was performed separately for sepsis at the levels of gene expression, methylation, and protein abundance to identify candidate genes associated with sepsis. These genes were validated using SMR analysis, and significant eQTLs and mQTLs were advanced to an integrated analysis followed by validation in an independent cohort. This rigorous process culminated in the identification of *CXCR2* as a notably associated gene (**Fig 1**).

**Fig 1.**
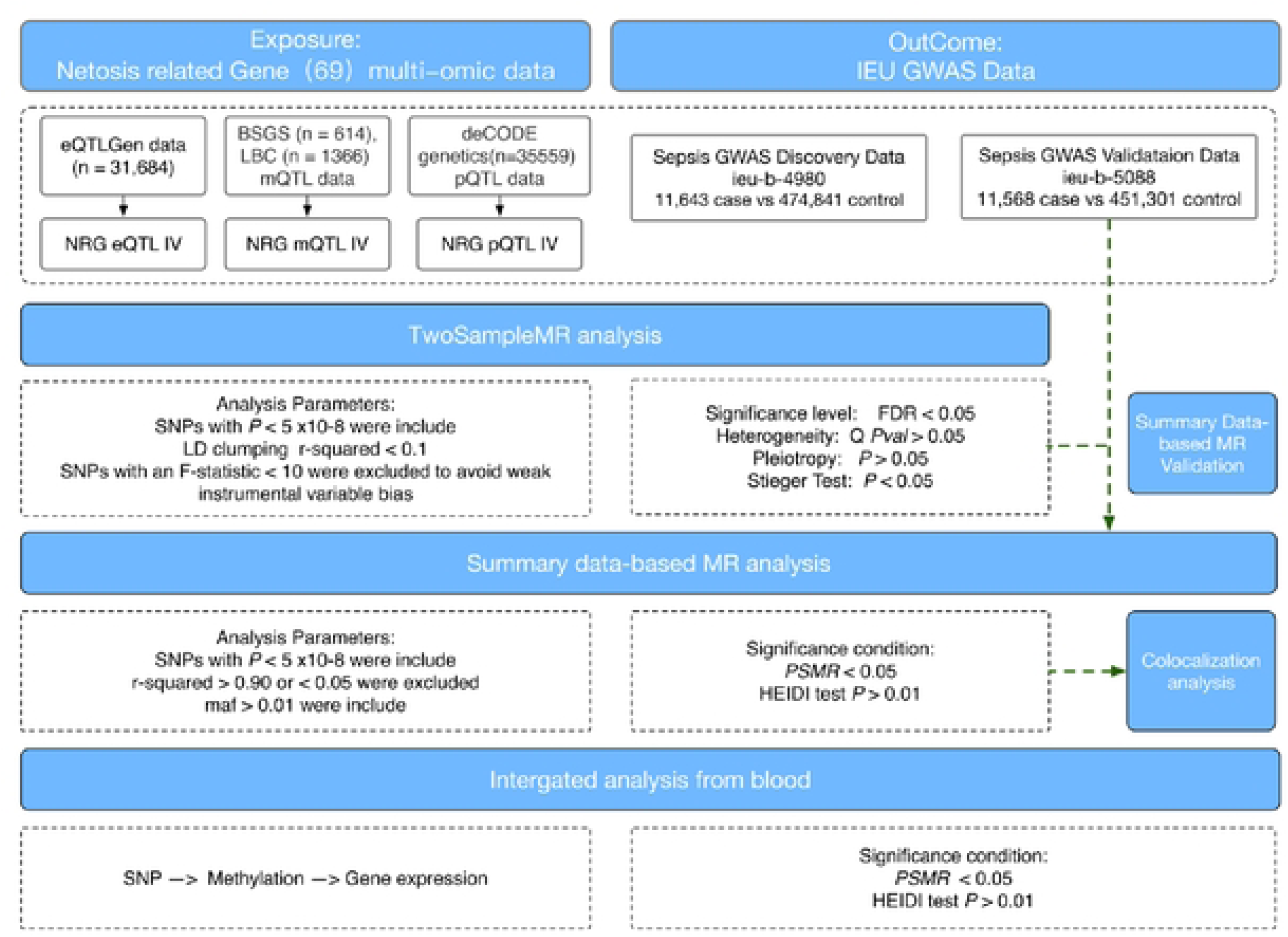
Overview of the study design.

### eQTL-based MR analysis

By intersecting our NRGs with key cis-eQTLs from the eQTLGen consortium, we identified 62 genes that met our criteria. Applying the established criteria for selecting IVs, we identified 438 cis-eQTLs associated with these 62 NRGs as the final IVs within our exposure data. A two-sample MR analysis of patients with sepsis was performed. Using the IVW method as our primary analytical approach, we identified four genes significantly associated with sepsis risk (FDR<0.05) (**S3 Table**, **Fig 2**). These genes include *CXCR1* (OR: 1.11, 95% CI: 1.04–1.17; FDR = 2.21 × 10^−2^), *CXCR2* (OR: 1.14, 95% CI: 1.07–1.21; FDR = 2.68 × 10^−3^), *ENTPD4* (OR: 1.16, 95% CI: 1.09–1.24; FDR = 2.49 × 10^−4^), and *MAPK3* (OR: 0.96, 95% CI: 0.93–0.99; FDR = 4.59 × 10^−2^). The primary analysis demonstrated the absence of heterogeneity (*P* > 0.05, **S4 Table**) and pleiotropy (*P* > 0.05, **S5 Table**). Additionally, Steiger filtering corroborated the directional relationship between outcomes and exposures (**S6 Table**).

**Fig 2.**
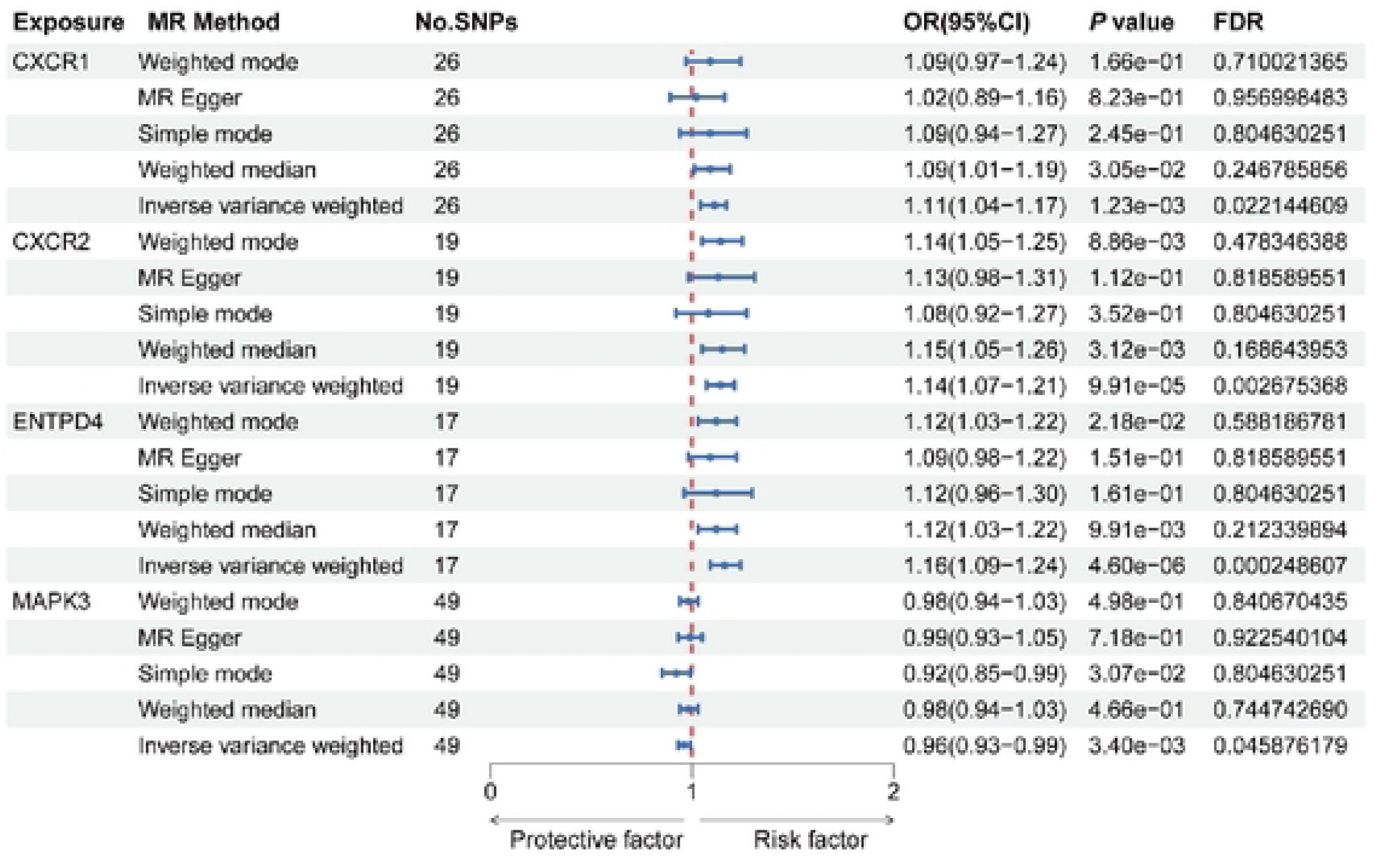
Mendelian randomization results for the association between the expression of neutrophil extracellular trap-related genes and sepsis risk.

### mQTL-based MR analysis

To further explore the epigenetic mechanisms influencing susceptibility to sepsis, we analyzed mQTLs associated with NRGs. Following stringent criteria for selecting IVs, we identified 780 cis-mQTLs across 55 NRGs that qualified as the final IVs within our exposure dataset. We then performed two-sample MR analysis of patients with sepsis. Using the IVW method as our primary analytical approach, we identified three CpG sites significantly associated with the sepsis risk (FDR<0.05) (**S7 Table**, **Fig 3**). These sites include cg06547715 (OR: 0.92, 95% CI: 0.88–0.97; FDR = 2.06 × 10^−2^), cg14150666 (OR: 0.95, 95% CI: 0.93–0.97; FDR = 3.46 × 10^−3^), and cg24126361 (OR: 0.97, 95% CI: 0.95–0.99; FDR = 2.06 × 10^−2^), associated with the genes *CXCR2* and *CXCR1*, respectively. The primary analysis did not indicate any evidence of heterogeneity (*P* > 0.05, **S8 Table**) or pleiotropy (*P* > 0.05, **S9 Table**).

**Fig 3.**
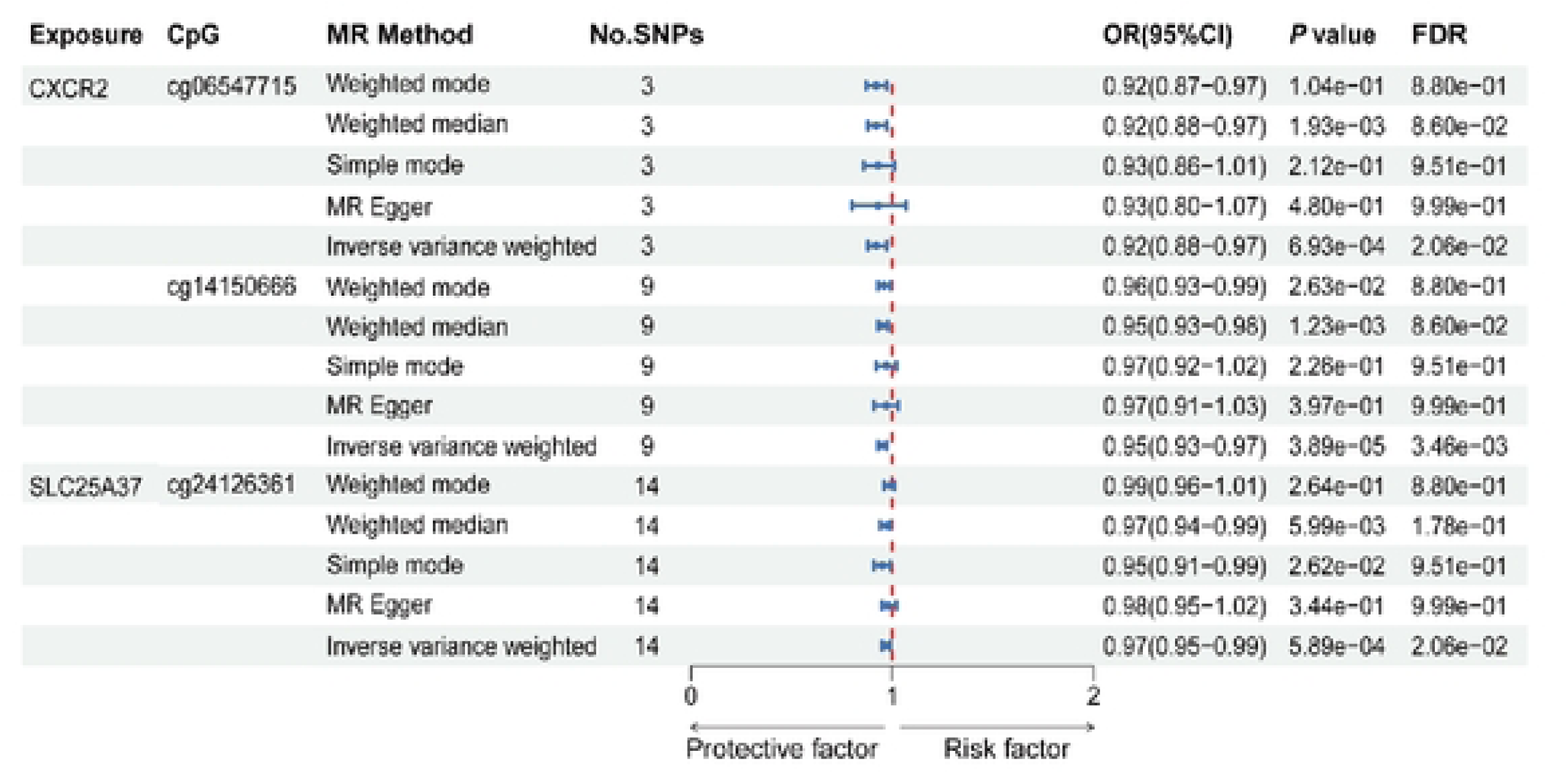
Mendelian randomization results for the association between neutrophil extracellular trap-related gene methylation and sepsis risk.

### pQTL-based MR analysis

By screening cis-pQTL data associated with NRGs, we identified 2304 SNPs across 36 NRGs in the exposure data as the final IVs based on the selection criteria (FDR < 0.05). Ten proteins were independently associated with sepsis risk. *SIGLEC14* (OR: 1.02, 95% CI: 1.01−1.04; FDR = 8.05 × 10^−3^) and *SIGLEC5* (OR: 1.02, 95% CI: 1.01−1.04; FDR = 1.13 × 10^−2^) were positively associated with sepsis risk. Conversely, *AKT2* (OR: 0.94, 95% CI: 0.90−0.97; FDR = 4.41 × 10^−3^), *CEACAM3* (OR: 0.75, 95% CI: 0.66−0.85; FDR = 7.57 × 10^−5^), *CTSG* (OR: 0.80, 95% CI: 0.69−0.92; FDR = 8.77 × 10^−3^), *F3* (OR: 0.70, 95% CI: 0.61−0.82; FDR = 7.57 × 10^−5^), *FCAR* (OR: 0.72, 95% CI: 0.56−0.91; FDR = 2.07 × 10^−2^), *HMGB1* (OR: 0.83, 95% CI: 0.76−0.90; FDR = 7.57 × 10^−5^), *IL17A* (OR: 0.63, 95% CI: 0.44−0.90; FDR = 3.55 × 10^−2^), and *ITGB2* (OR: 0.77, 95% CI: 0.69−0.88; FDR = 3.19 × 10^−4^), were inversely associated with sepsis risk (**S10 Table**, **Fig 4**). In the primary analysis, no heterogeneity (*P* > 0.05, **S11 Table**) or horizontal pleiotropy (*P* > 0.05, **S12 Table**) was observed.

**Fig 4.**
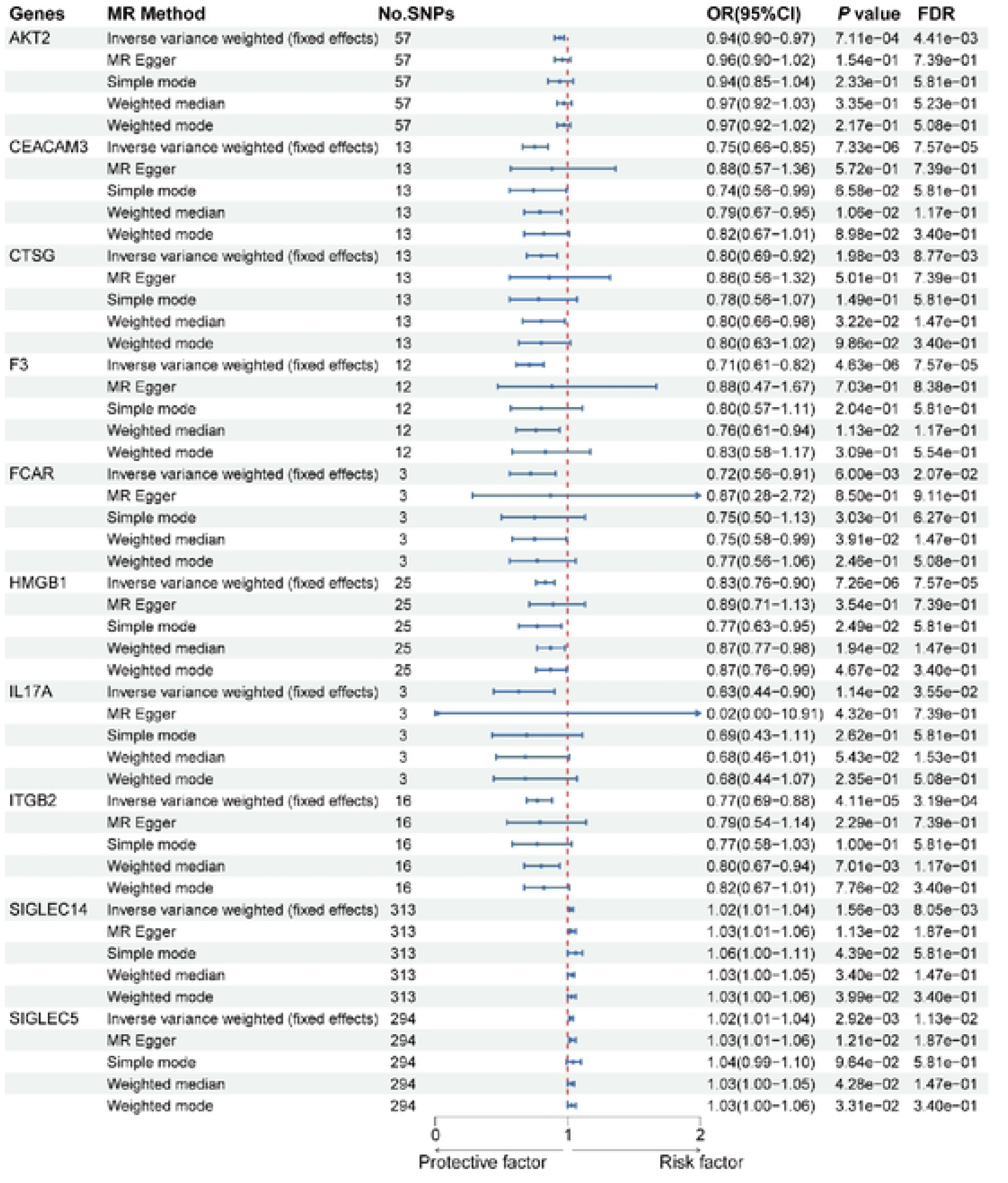
Mendelian randomization results for the association between neutrophil extracellular trap-related proteins and sepsis risk.

### SMR and colocalization analyses

Further SMR analysis was conducted to verify the causal relationships between mQTLs and sepsis, eQTLs and sepsis, as well as pQTLs and sepsis. The protein data were not significant after SMR analysis, and the *CXCR2* gene was identified through mQTL and eQTL SMR analyses. The results indicated that *CXCR2* was consistent with the direction of the effect observed in the MR analysis and passed the HEIDI (*P* > 0.01) and SMR tests (*P* < 0.05), suggesting a causal relationship unconfounded by pleiotropy. The SMR plots and effect estimates are presented in the **S13 Table** and **Fig 5A**, **B**. Other genes did not demonstrate significant associations in these analyses. Upon further exploring the causal relationships between these genes and sepsis, colocalization analysis revealed the details of SNPs (**Fig 5B**) and showed that *CXCR2* (OR: 1.16, 95% CI: 1.05−1.29; PPH4 = 1), cg06547715 (OR: 0.92, 95% CI: 0.87−0.97; PPH4 = 0.912), and cg14150666 (OR: 0.96, 95% CI: 0.93−0.99; PPH4 = 1) shared a causal variant with sepsis, which added another evidence to our results (**S14-S16 Tables**, **Fig 5C**).

**Fig 5.**
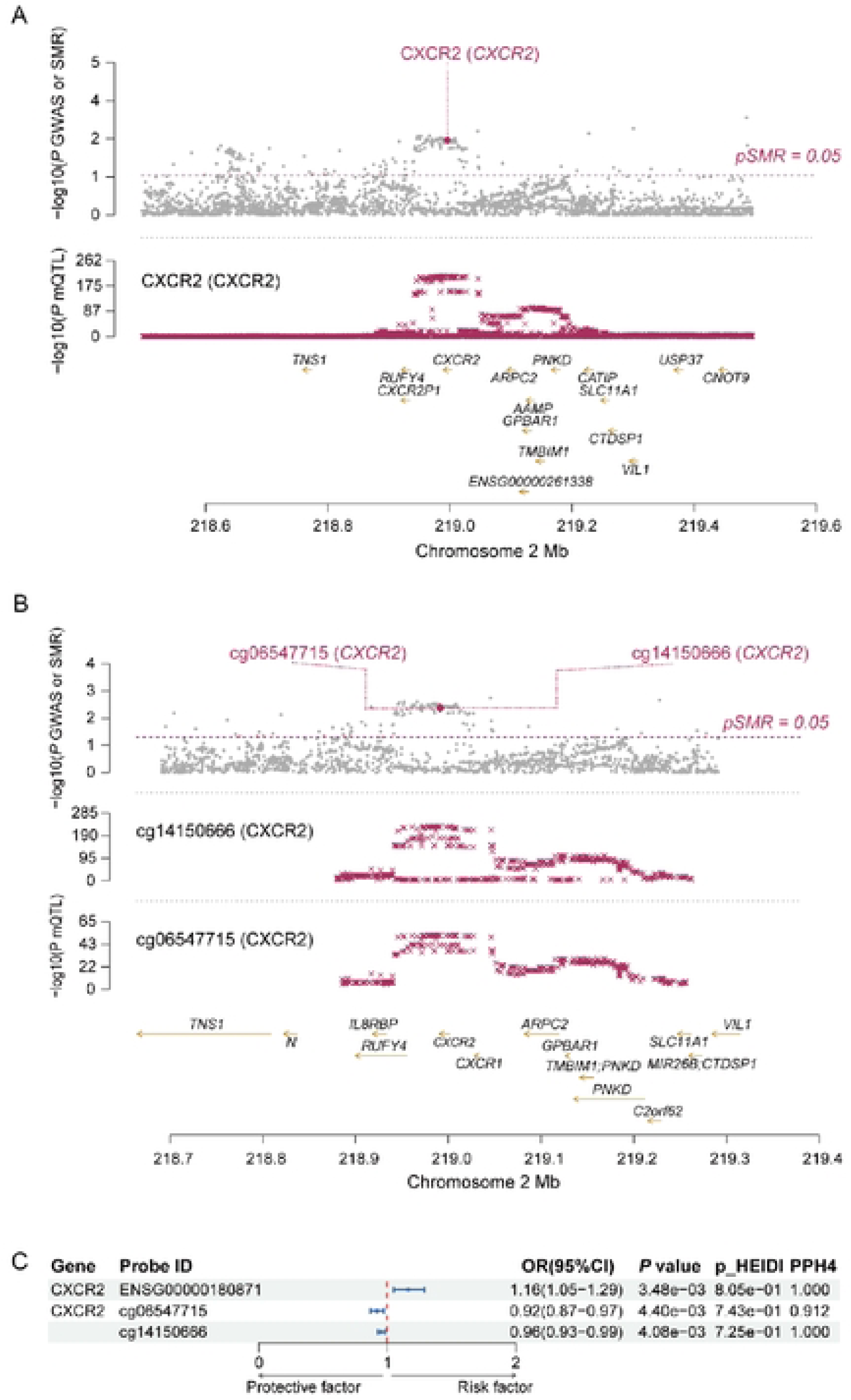
SMR and colocalization analysis results in discovery datasets (ieu-b-4980 cohort). A. MR association between *CXCR2* expression and sepsis risk. B. MR association between *CXCR2* methylation and sepsis risk. C. Colocalization evidence of the expression and CpG site methylation of *CXCR2* and sepsis susceptibility.

### Integration of GWAS and *CXCR2*-related eQTL/mQTL data from the blood

Subsequently, we hypothesized that gene expression could be the underlying mechanism explaining the plausible causality in the disease. Moreover, DNA methylation (DNAm) in promoters or enhancers typically plays a crucial role in the regulation of disease-associated target genes. In this study, we identified *CXCR2*, which encodes a protein belonging to the G protein-coupled receptor family. This protein serves as a receptor for interleukin 8, which binds with high affinity and transmits signals through the G-protein-activated second messenger system. Additionally, this receptor can bind to chemokine (C-X-C motif) ligand 1 (*CXCL1/MGSA*), which is known to stimulate melanoma growth. This receptor mediates the migration of neutrophils to sites of inflammation. After three-step SMR analysis, we discovered that the DNAm probe cg06547715 located in the enhancer region of *CXCR2* plays a crucial role in gene expression regulation. Methylation levels at this site negatively correlate with *CXCR2* expression (βSMR = −0.658) and sepsis (βSMR = −0.082). Conversely, the expression levels of *CXCR2* are positively correlated with sepsis (βSMR = 0.13). This suggests that reduced DNAm levels within the enhancer region of *CXCR2* may upregulate its expression, increasing susceptibility to sepsis (**S17 Table**).

### Replication of results in other cohorts

We performed SMR analysis of *CXCR2* in the ieu-b-5088 cohort to validate our findings. The results showed the methylation level at the cg06547715 site negatively correlated with sepsis (βSMR = −0.0604334), and *CXCR2* expression levels were positively correlated with the condition (βSMR = 0.114256), aligning with the effects observed in cohort ieu-b-4980 (**S18 Table**; **Fig 6A**, **B)**. Colocalization analysis depicted the details of SNPs locus (**Fig 5B**) and showed that *CXCR2* (OR: 1.12, 95% CI: 1.01−1.24; PPH4=1), cg06547715 (OR: 0.94, 95% CI: 0.89−0.99; PPH4=0.912), and cg14150666 (OR 0.97, 95% CI 0.94−1.00; PPH4=1) shared a causal variant with sepsis, which added additional evidence to our results (**S19-S21 Tables**; **Fig 6C**).

**Fig 6.**
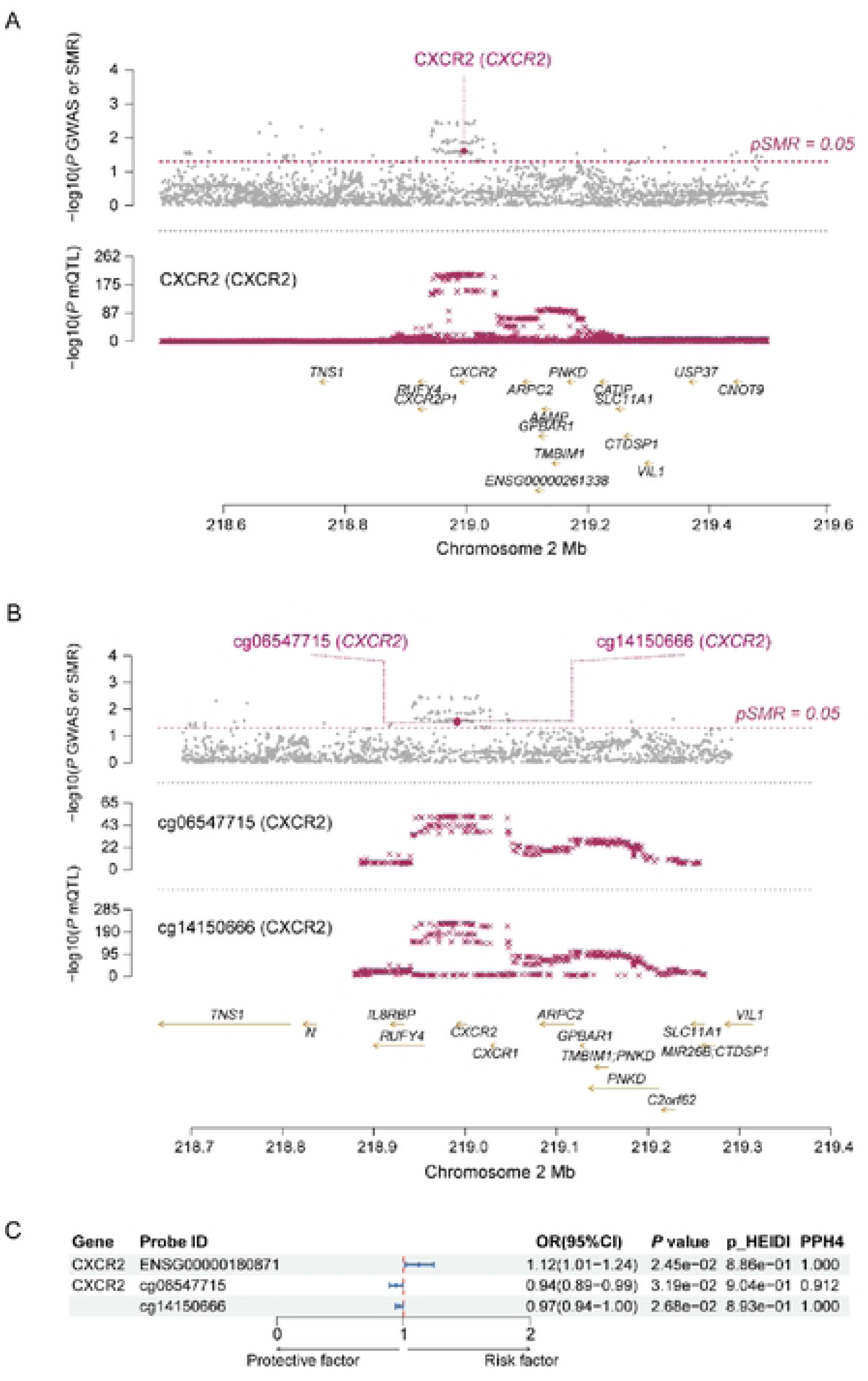
SMR and colocalization analysis results in validation datasets (ieu-b-5088 cohort). A. MR association between *CXCR2* expression and sepsis risk. B. MR association between *CXCR2* methylation and sepsis risk. C. Colocalization evidence of the expression and CpG site methylation of *CXCR2* and susceptibility to sepsis.

## Discussion

This study leveraged the multiomic MR method to explore the relationship of NRGs with sepsis risk. Our findings not only contribute to the existing literature but also provide a foundation for future therapeutic interventions. The identification of *CXCR1*, *CXCR2*, *ENTPD4*, and *MAPK3* as genes significantly associated with sepsis risk is consistent with the growing body of evidence suggesting that chemokine signaling and immune response genes are critical in sepsis pathogenesis. The involvement of *CXCR1* underscores the critical role of neutrophil chemotaxis in sepsis. Our findings are consistent with those of previous studies, suggesting that polymorphisms in these genes modulate the immune response to infection[29, 30]. The association between *ENTPD4* and sepsis risk is intriguing, given its role in purinergic signaling, which has been implicated in the regulation of NET formation[31, 32]. Similarly, the observed association with *MAPK3*, a key regulator of cellular stress responses, aligns with the hypothesis that NETs are part of a broader inflammatory network during sepsis[33, 34]. The robust association of *CXCR2*, in particular, resonates with prior research indicating its role in modulating neutrophil migration and the inflammatory cascade central to sepsis[35–37]. Our multiomic MR approach, which integrates genetic, epigenetic, and proteomic data, strengthens the causal inference by mitigating confounding and reverse causation biases inherent in observational studies. The results of colocalization analysis provided us with additional evidence supporting the role of the *CXCR2* gene in sepsis. Our analysis showed that the gene expression of *CXCR2* is significantly associated with the risk of sepsis development, and the PPH4 value is greater than 0.75, indicating that the association between GWAS and eQTL may be caused by the same causal variant. This finding further strengthens the possibility of using *CXCR2* as a potential therapeutic target for sepsis.

The inverse correlation between DNAm at cg06547715 and *CXCR2* expression and its subsequent impact on sepsis risk offer a novel perspective on the epigenetic modulation of immune response genes[38, 39]. This aligns with the emerging paradigm that epigenetic modifications, especially DNAm, are dynamic and responsive to environmental stimuli, potentially serving as a molecular switch in the context of sepsis[40]. The implications of our findings extend to the realm of therapeutics, where targeting *CXCR2* and its regulatory mechanisms could represent a new frontier in sepsis management[30]. The potential to modulate gene expression through epigenetic drugs or gene therapies offers a promising avenue for mitigating the hyperinflammatory state characteristic of sepsis. Furthermore, the pQTL analysis revealed a direct link between specific proteins and the risk of sepsis development. By screening cis-pQTL data associated with NRGs, we identified 10 proteins that are significantly associated with the risk of sepsis. For instance, *SIGLEC14* and *SIGLEC5* are positively correlated, whereas *AKT2*, *CEACAM3*, *CTSG*, *F3*, *FCAR*, *HMGB1*, *IL17A*, and *ITGB2* are negatively correlated with the risk of sepsis development. These findings suggest that these proteins may play a key role in the pathogenesis of sepsis, offering new leads for the development of targeted therapeutic strategies in the future[41–46]. Our results are in line with those of previous studies that have implicated NRGs in various inflammatory and autoimmune conditions, suggesting a common pathway in the dysregulation of immune responses[47, 48]. The consistency of our findings with those from independent cohorts and the replication of results across different analytical approaches lend credence to the robustness of our conclusions. While our multiomic MR approach provides a robust platform for causal inference, it is not without limitations. The reliance on observational data exposes our study to potential unmeasured confounding despite the use of MR methods to control for such biases. Due to the limited number of NRG proteins in the pQTL dataset, the present study did not fully explore the causal relationship between mitochondrial proteins and the risk of sepsis development.

Future research should aim to replicate our findings in diverse populations and investigate the functional consequences of the genetic and epigenetic variants identified. Additionally, experimental studies are warranted to elucidate the mechanistic links among NRGs, inflammation, and sepsis development, potentially leading to the discovery of new biomarkers and therapeutic targets.

In conclusion, our multiomic MR study provides a deeper understanding of the role of NRGs in sepsis, with a particular focus on *CXCR2*. Our findings have significant implications for the development of precision medicine strategies for sepsis and highlight the importance of integrating genetic and epigenetic data in the quest for novel therapeutic targets. The identification of *CXCR2* as a key gene in sepsis risk offers a promising starting point for future research and interventional studies aimed at improving the outcomes in patients with this devastating condition.

## Data Availability

All data produced in the present work are contained in the manuscript

https://kdocs.cn/l/cugdp5seocwJ

## Acknowledgments

We gratefully acknowledge the authors and participants of all GWASs, including the IEU OpenGWAS Project, eQTLgen, the mQTL summary data of McRae et al., and the deCODE Genetics dataset, from which we utilized summary statistics data.

## Notes

### Competing Interest Statement

The authors have declared that no competing interests exist.

### Clinical Protocols

https://kdocs.cn/l/cugdp5seocwJ

### Funding Statement

This study did not receive any funding

### Author Declarations

The study utilized publicly available genomic data from sources such as the "IEU OpenGWAS Project" database, eQTLGen Consortium, mQTL summary data, and deCODE Genetics dataset.

